# A Quantitative Framework to Study Potential Benefits and Harms of Multi-cancer Early Detection Testing

**DOI:** 10.1101/2021.03.19.21253904

**Authors:** Boshen Jiao, Roman Gulati, Hormuzd A. Katki, Philip E. Castle, Ruth Etzioni

## Abstract

**Background:** Multi-cancer tests permit identification of multiple cancers with one blood draw. The objective of this study was to quantify potential population impact of a multi-cancer test.

**Methods:** We formulate mathematical expressions for expected numbers of (1) individuals exposed to unnecessary confirmation tests (*UCT*), (2) cancers detected (*CD*), and (3) lives saved (*LS*) given disease prevalence and mortality and the test’s performance and expected mortality reduction. We consider additions of colorectal, liver, lung, ovary, and pancreatic cancer to a test for breast cancer using published performance characteristics of a multi-cancer test and prevalence of each cancer at ages 50, 60, or 70 based on 5-year incidence rates and corresponding 15-year probabilities of cancer death in the Surveillance, Epidemiology, and End Results registry, assuming 20% mortality reduction for each.

**Results:** *UCT* depends on screening age but is overwhelmingly determined by overall specificity of the test and is relatively insensitive to the types and number of cancers included. For a given overall specificity, *UCT/CD* is most favorable for higher-prevalence cancers (e.g., *UCT*/*CD* = 5.6 for breast+lung versus 6.5 for breast+liver at age 50). Under a common mortality reduction, *UCT*/*LS* is most favorable when the test includes higher-mortality cancers (e.g., *UCT*/*LS* = 48.5 for breast+lung versus 74.7 for breast+liver at age 50).

**Conclusions:** The harm-benefit tradeoffs of multi-cancer testing depend on the number and type of cancers included. Overall specificity is paramount for controlling unnecessary confirmation tests. For a given overall specificity, multi-cancer tests should prioritize prevalent and/or lethal cancers for which curative treatments exist.

---

The advent of liquid biopsy technology has ushered in a new era of cancer diagnostics. Assays to detect circulating cell-free DNA supplemented by protein and methylation signatures promise to dramatically alter the landscape of cancer surveillance and early detection. In particular, the possibility of multi-cancer early detection, where a single blood sample is interrogated for multiple cancers, is attracting a great deal of attention.^1,2^

Several multi-cancer tests are in development, each harnessing different features of the circulating tumor DNA. Liu et al present a test using targeted methylation analysis of circulating cell-free DNA that in principle detects and localizes more than 50 cancer types, and quantify its sensitivity for a pre-specified set of 12 cancers.^3^ Cohen et al present diagnostic performance of a test using circulating DNA and protein biomarkers;^4^ Lennon et al identify 10 cancer types using an updated version of the test.^5^ And Cristiano et al present a test that uses fragmentation patterns of cell-free DNA across the genome along with mutation-based cell-free DNA, estimating the diagnostic performance of this test across 7 cancer types.^6^

Multi-cancer testing offers the potential for improved diagnosis of cancers where tests already exist, such as mammograms in women with dense breasts. It is less invasive than some existing tests, such as colonoscopies for colorectal cancer. Further, multi-cancer tests include cancers for which tests do not currently exist, possibly because the search for biomarkers with adequate sensitivity and specificity has been unsuccessful. For other cancers, it is simply not practical to deploy individual screening tests in the general population due to extremely low prevalence.^7^ Aggregating diagnosis of these cancers into a single test could increase the combined prevalence to an acceptable level for population screening. There may also be an economic advantage to doing a single test rather than a series of individual tests.

At this point however, many questions exist about the likely impact of multi-cancer testing on population outcome including benefits, such as cancer deaths prevented, and harms, such as unnecessary imaging tests or biopsies.^1^ These outcomes depend critically on disease prevalence at the time of the test and mortality in the absence of the test in addition to test performance, particularly its ability to detect and treat potentially fatal disease early.

This study investigates how the potential benefits and harms of a multi-cancer test depend on test and disease characteristics. We first explore the simple setting of hypothetical two-cancer tests and then examine realistic tests involving up to six cancers. Our analysis creates a quantitative framework to project the population impact of multi-cancer tests while pointing to criteria for the number and the type of cancers to include.

## Methods

### Overview

Suppose that we have a test for a single cancer (cancer *A*) and our goal is to evaluate a test that includes two cancers (cancers *A* and *B*). We assume that the test produces an assessment of whether cancer is present and indicates a tissue of origin (TOO). We further assume that individuals do not have both cancers present concurrently at the time of the test. We present results for hypothetical two-cancer tests and show how they depend on test performance and cancer characteristics. We then present results for breast cancer (cancer *A*) and colorectal, liver, lung, ovary, or pancreatic cancer (cancer *B*), evaluating the harm-benefit tradeoffs as we sequentially build up to six cancers based on published characteristics of an existing multi-cancer test. We produce results for single-occasion testing at ages 50, 60, or 70 to explore age dependence of the outcomes.

### Test performance

We distinguish between the *sensitivity of a single-cancer test*, which is the probability that the test will return a positive result if the cancer is present, and the *sensitivity of a multi-cancer test*, which has two components: (1) the *overall sensitivity*, which is the probability the test returns a cancer signature given that a targeted cancer is present and (2) the *probability of correct localization* for each targeted cancer. For cancer *A*, this is the probability that the test returns a cancer signature and identifies cancer *A* when it is present. We note that, while our discussion refers to sensitivity in general terms, it is the sensitivity to detect early-stage tumors that counts. Published multi-cancer studies have shown considerably poorer sensitivities for early compared with advanced-stage tumors.^4^

We define the *marginal sensitivity of a multi-cancer test* as the probability that the multi-cancer test identifies a specific cancer when it is present. The marginal sensitivity for cancer *A* can be written:

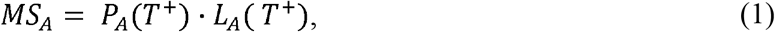

where *P_A_* (*T*+) is the overall sensitivity of the multi-cancer test, i.e., the probability the test returns a cancer signal given cancer *A* is present, and *L_A_*(*T*^+^) is the probability of correctly localizing the TOO as *A* given that the multi-cancer test returns a cancer signal (*T*+) and cancer *A* is in fact present. With *k* targeted cancers there are *k* marginal sensitivities.

We define the *specificity of a multi-cancer test* as the probability the test returns a non-cancer signature when none of the targeted cancers is present.

### Outcome metrics

In single-cancer testing, a key metric used to evaluate harm-benefit is the positive predictive value (*PPV*). In this setting, 1/*PPV* is often cited as the number of biopsies required per cancer detected, and 1/*PPV* - 1 the number of unnecessary biopsies per cancer detected, assuming that all positive tests are followed by a biopsy.

In single-cancer testing, each individual is subject to only at most one biopsy, so the unnecessary biopsies arise solely from false positive tests, i.e., from individuals without cancer. In multi-cancer testing, however, unnecessary confirmation tests may occur in individuals who have one of the cancers included in the test. This could happen when the multi-cancer test correctly returns a cancer signal but incorrectly identifies the TOO. In this case, a confirmation test performed to verify that cancer is present in the putative TOO would constitute an unnecessary confirmation test. Thus, in multi-cancer testing, unnecessary confirmation tests arise from individuals without any of the targeted cancers as well as from those whose tumor is incorrectly localized.

For a two-cancer test, we can directly formulate the expected number of screened individuals potentially exposed to unnecessary confirmation tests as:

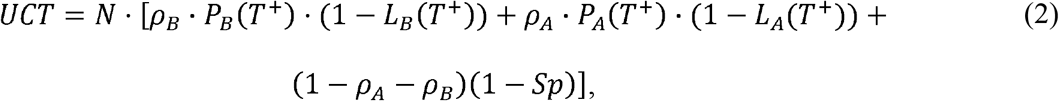

where *p_A_* and *p_B_* are the prevalence of cancers *A* and *B*, respectively, and *S_p_* is the overall specificity of the test. The first two terms in square brackets reflect the correct return of a cancer signal but incorrect localization, and the third term reflects a false positive. In practice, if the overall specificity is even modestly below 100% and the prevalence of each cancer is not high, the third term will dominate.

We follow common practice in single-cancer testing and normalize *UCT* by the expected number of cancers detected. This depends on the marginal sensitivity of the multi-cancer test to detect each cancer and, in the setting of two cancers, is given by:

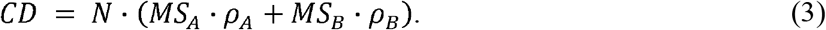

The formulation of *CD* reflects the expected number of people diagnosed with cancer following confirmation precipitated by the multi-cancer test. In practice, both *UCT* and *CD* will depend on the protocol for further confirmation testing in the event that confirmation testing of the putative TOO returns a negative result.

In addition to *UCT* and *CD*, we project the expected lives saved (*LS*) by assuming a disease-specific mortality reduction for each targeted cancer and applying this value to the cumulative risk of disease-specific death without screening. The mortality reduction corresponding to any screening test is a complex function of the interaction between screening test performance, screening protocol, and disease natural history. Rather than explicitly modeling these interactions for each cancer, we assume a value for the mortality reduction based on published breast cancer screening trials.^11^ The expected number of lives saved is given by:

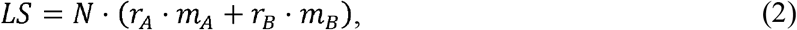

where *r_A_* and *r_B_* are the mortality reductions for cancers *A* and *B*, respectively, and mA and mB are corresponding probabilities of cancer-specific death. In addition to *UCT*/*CD*, we also examine *UCT*/*LS* as a harm-benefit measure. General expressions for the outcome metrics for a *k*-cancer test are given in the **Supplementary Appendix** (available online).

### Evaluation of hypothetical and realistic multi-cancer tests

We first consider a two-cancer test that screens for (*A*) breast cancer and (*B*) colorectal, liver, lung, ovary, or pancreatic cancer among women in the United States. We base sensitivities and correct localization probabilities on estimates from Liu et al for cancers in stages I-III,^3,9^ and overall specificity on their specificity estimates. Age-specific prevalence of cancer is based on incidence rates observed in the Surveillance, Epidemiology, and End Results (SEER) registry over the years 2000-2002 inclusive.^8^ We assume that cancers diagnosed within the 5-year age groups 50-54, 60-64, and 70-74 years are prevalent and thus could potentially be detected by a multi-cancer test given at ages 50, 60, and 70 years, respectively.

At any given age, screening cannot affect mortality among cancers diagnosed prior to that age. Therefore, we use SEER incidence-based mortality, which permits calculation of cumulative disease-specific mortality among cancers diagnosed after the specified screening age.^12^ We estimate 15-year incidence-based mortality and apply the assumed mortality reduction to project expected lives saved. We restrict the time interval for mortality to 10 years in a sensitivity analysis.

For screening benefit we assume a 20% mortality reduction, which corresponds to the consensus benefit estimate across breast cancer screening trials.^11^ In doing so, we project outcomes were multi-cancer screening as beneficial on a per-cancer basis as in these trials. Since we are modeling only single-occasion testing, we examine how our outcome metrics change under a more modest benefit (5% mortality reduction per cancer). Moreover, in a second sensitivity analysis, we use a 10% mortality reduction for the three high-prevalence cancers for which screening tests currently exist and a 50% mortality reduction for the three low-prevalence cancers.

## Results

### A hypothetical two-cancer test

Figure 1. shows *UCT* and *CD* for a hypothetical two-cancer test under specified diagnostic characteristics and prevalence settings. The values used for prevalence of cancer *B* are low in keeping with the estimates derived from the SEER registry examined below. Projected values for a wider range of settings are shown in **Supplementary Table 1** (available online).

**Figure 1.**
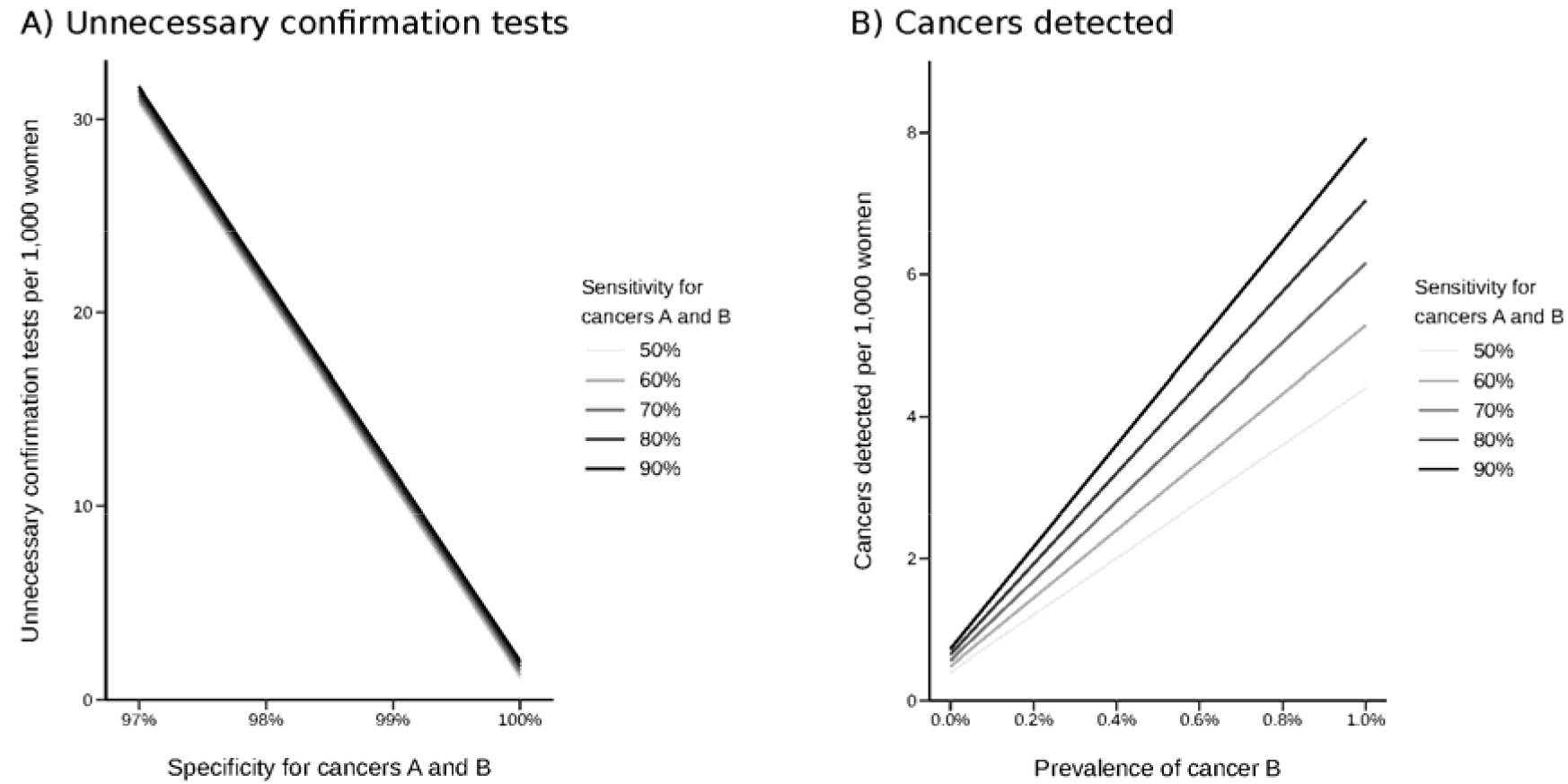
Hypothetical outcomes of a two-cancer test per 1,000 individuals: A) expected number of individuals potentially exposed to unnecessary confirmation tests given sensitivity and specificity for both cancers and B) expected cancers detected given prevalence of cancer B and sensitivity for both cancers assuming the prevalence of cancer A is 0.1%.

Results show that *UCT* is driven primarily by the specificity of the test. Its dependence on sensitivity is small; its dependence on prevalence and localization accuracy (not shown) are similar in magnitude. *CD* increases with the prevalence of cancer *B* and the sensitivity of the test, leading to lower *UCT*/*CD* for higher-prevalence cancers. While the results are intuitive, the message is that benefit-harm tradeoffs of multi-cancer testing depend on multiple factors pertaining to characteristics of the test and the cancers included.

### A realistic six-cancer test

**Table 1** gives age-specific 5-year risks of cancer diagnosis, which we use to approximate prevalence, and risks of cancer death for the cancers considered. As expected, breast and lung cancer have the highest prevalence while liver cancer has the lowest. All prevalence estimates are less than 0.5%, and they are less than 0.1% for liver, ovary, pancreatic cancers. **Table 2** summarizes overall sensitivities and localization probabilities from Liu et al^10^ for these cancers. The highest marginal sensitivity is for colorectal cancer; the lowest is for liver cancer.

**Table 1.** Five-year net cumulative incidence and 15-year incidence-based mortality for selected cancers based on diagnoses in the Surveillance, Epidemiology, and End Results program in 2000-2002.

| Tissue of origin | Age, y | 5-year probability of diagnosis, % | 15-year probability of death, % |
| --- | --- | --- | --- |
| Breast | 50 | 0.249 | 0.065 |
|  | 60 | 0.379 | 0.132 |
|  | 70 | 0.447 | 0.259 |
| Colorectal | 50 | 0.047 | 0.019 |
|  | 60 | 0.104 | 0.053 |
|  | 70 | 0.208 | 0.141 |
| Lung | 50 | 0.049 | 0.039 |
|  | 60 | 0.166 | 0.141 |
|  | 70 | 0.305 | 0.270 |
| Ovary | 50 | 0.023 | 0.014 |
|  | 60 | 0.040 | 0.030 |
|  | 70 | 0.051 | 0.043 |
| Pancreas | 50 | 0.008 | 0.007 |
|  | 60 | 0.023 | 0.021 |
|  | 70 | 0.052 | 0.048 |
| Liver | 50 | 0.003 | 0.003 |
|  | 60 | 0.009 | 0.008 |
|  | 70 | 0.017 | 0.016 |

**Table 2.**
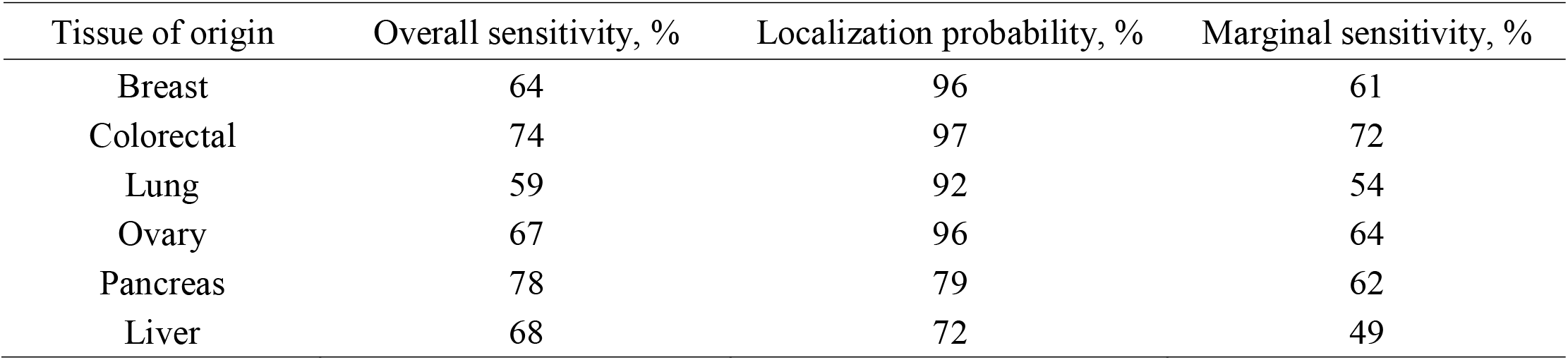
Overall sensitivity, localization probability, and implied marginal sensitivity of multi-cancer test for selected stage I-III cancers based on Liu et al.

The 5 two-cancer tests (breast cancer plus one of the other cancers) are associated with similar *UCT* (**Supplementary Table 2**), but they show clear patterns of higher *UCT*/*CD* when prevalence of the candidate second cancer is lower (**Figure 2**). Assuming similar mortality reductions across cancers, *UCT*/*LS* is an order of magnitude greater than *UCT*/*CD*. Both harm-to-benefit measures improve with screening age and are most favorable when the second cancer is lung cancer, which has the highest prevalence and mortality, except at age 50 years when *UCT*/*CD* is most favorable for breast+colorectal cancer. These features lead to lung cancer having the highest *CD* and *LS* at ages 60 and 70 despite the marginal sensitivity for lung cancer being relatively low. Based on these metrics, the test for breast+lung cancer is optimal among the two-cancer tests considered at these ages. Similar patterns are observed under a more modest screening benefit; **Figure 2** and **Supplementary Table 2** also show how *UCT*/*LS* changes when the screening benefit is 5% versus 20%.

**Figure 2.**
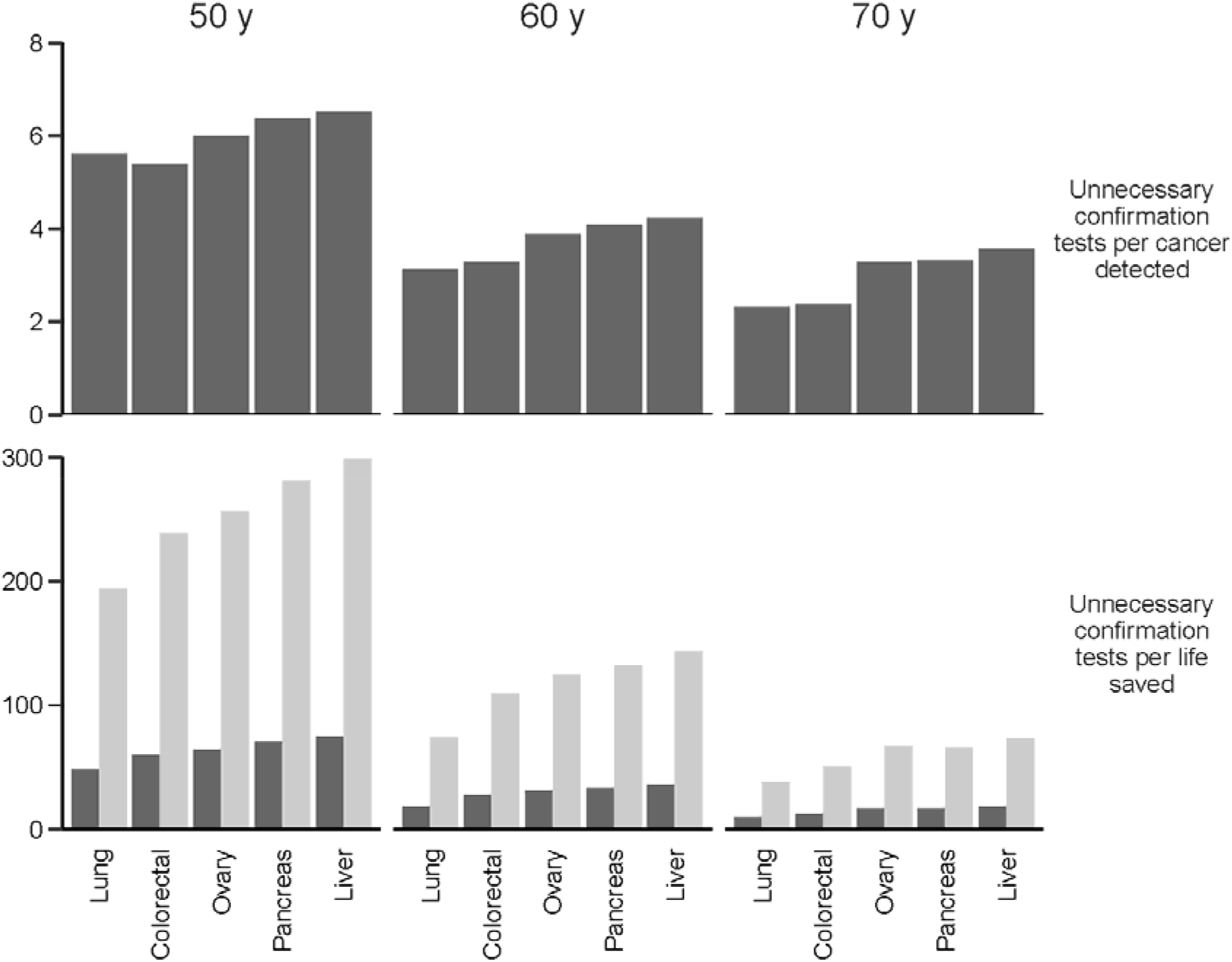
Unnecessary confirmation tests per cancer detected (top row) and per life saved (bottom row) associated with adding specific cancers to an existing test for breast cancer at specific screening ages (columns) assuming specificity of the two-cancer test is 99%, 15-year incidence-based mortality from the Surveillance, Epidemiology, and End Results program, and the mortality reduction for each cancer is 5% (light bars) or 20% (dark bars).

Similar patterns are observed when adding a third cancer to a breast+lung cancer test (**Figure 3** and **Supplementary Table 3**). Once again, *UCT* is similar across tests considered, and the optimal candidate has the highest prevalence and mortality, in this case colorectal cancer. Building up to a six-cancer test in this way yields the projected outcomes shown in **Table 3**. For comparison purposes, outcomes are presented also for a single-cancer test for breast cancer.

**Table 3.** Expected number of women potentially exposed to unnecessary confirmation tests (*UCT*), expected cancers detected (*CD*), expected lives saved (*LS*), and harm-benefit ratios *UCT*/*EC* and *UCT*/*LS*. Table shows results for a multi-cancer test for breast, lung, colorectal, ovarian, pancreatic, and liver cancers, and a single-cancer test for breast cancer with equal marginal sensitivity and specificity (99%). Specificity is 99% and mortality reduction for each cancer is 20%.

| Test | Screening age, y | Unnecessary confirmation tests, n | Cancers detected, n | Lives saved, n | UCT/CD | UCT/LS |
| --- | --- | --- | --- | --- | --- | --- |
| Multi-cancer | 50 | 10.1 | 2.3 | 0.3 | 4.3 | 34.5 |
|  | 60 | 10.2 | 4.4 | 0.8 | 2.3 | 13.3 |
|  | 70 | 10.3 | 6.6 | 1.6 | 1.6 | 6.7 |
| Single cancer | 50 | 10.0 | 1.5 | 0.1 | 6.6 | 77.8 |
|  | 60 | 10.1 | 2.3 | 0.3 | 4.3 | 38.2 |
|  | 70 | 10.1 | 2.7 | 0.5 | 3.7 | 19.5 |

**Figure 3.**
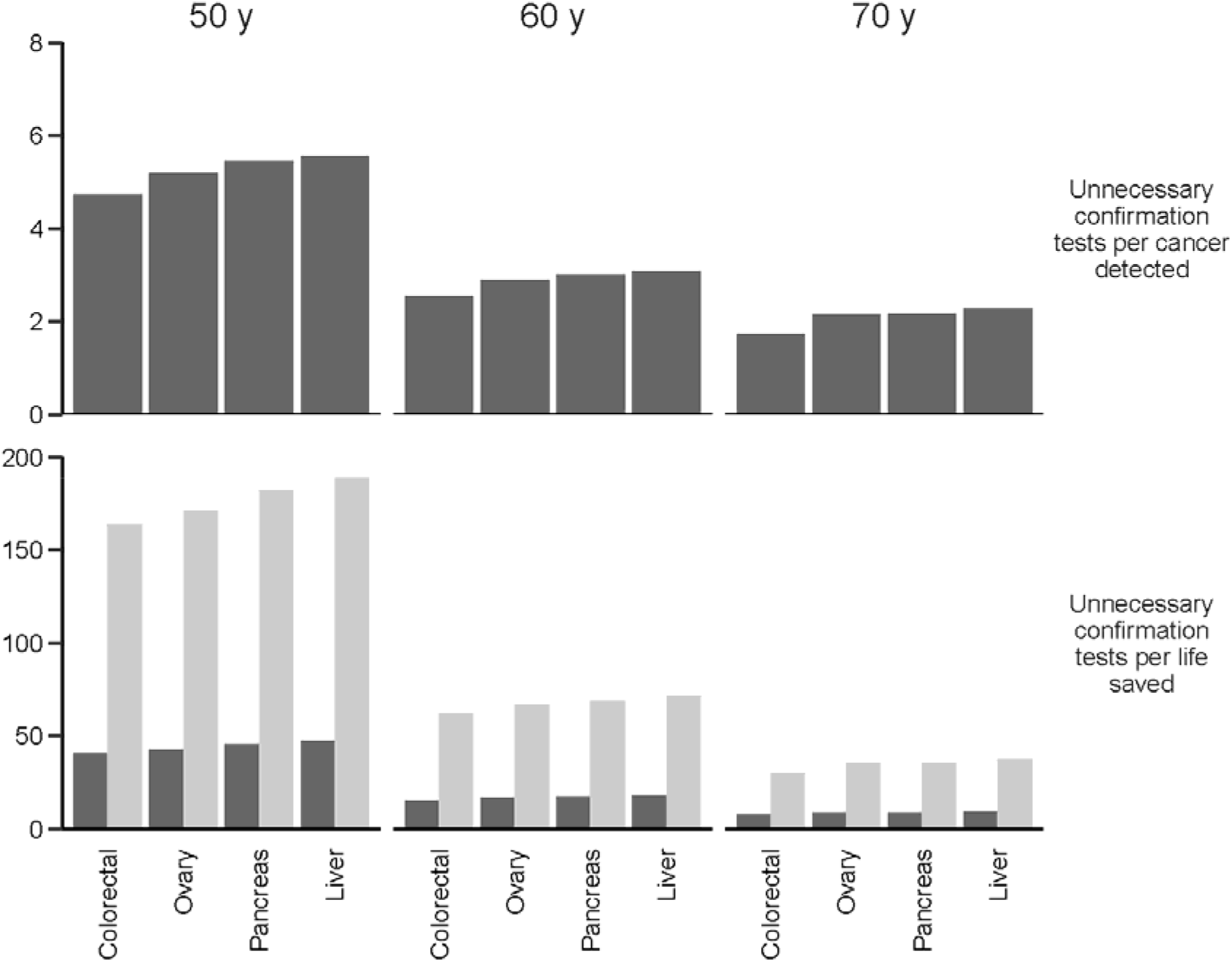
Unnecessary confirmation tests per cancer detected (top row) and per life saved (bottom row) associated with adding specific cancers to an existing test for breast and lung cancers at specific screening ages (columns) assuming specificity of the three-cancer test is 99%, 15-year incidence-based mortality from the Surveillance, Epidemiology, and End Results program, and the mortality reduction for each cancer is 5% (light bars) or 20% (dark bars).

Improving the mortality reduction for low-prevalence cancers from 20% to 50% and attenuating the mortality reduction from 20% to 10% for high-prevalence cancers alters the ordering of the two-cancer tests in terms of benefit-harm tradeoffs. Under these settings, the breast+ovary two-cancer test is associated with lower *UCT*/*LS* than the breast+lung cancer test for 50- and 60-year-old women (**Supplementary Figure 1**). Restricting attention from 15-to 10-year probabilities of cancer death (**Supplementary Figure 2**) does not alter the original patterns in the benefit-harm measures. Based on these measures, the same optimal sequence of cancers would be added to a multi-cancer test provided the absolute tradeoffs are judged to be acceptable.

## Discussion

In this study we present a quantitative framework for studying the potential impact of novel multi-cancer screening tests on population outcomes.Ultimately, both prevalence of and mortality from the included cancers play into harm-benefit tradeoffs that reflect population impact.

The need for a framework to clarify the multi-cancer testing outcomes is necessitated by the ways in which this new technology alters concepts of foundational diagnostic metrics. In addition to multiple concepts of test sensitivity, we must reconsider the standard definition of specificity in the setting of a multi-cancer test. In single-cancer testing, a truly negative test is taken to imply that the individual being tested does not have cancer. In multi-cancer testing for a specified set of cancers, our definition of specificity would mean that a truly negative test implies that the person being tested does not have one of the cancers in the targeted set.

Our analysis considers a limited set of harm-benefit metrics based on information currently available. More precise projection of any of these metrics would require information about disease natural history and screening test performance in a prospective setting. This information would be also needed to explore other metrics of benefit and harm, such as overdiagnosis, and to extrapolate from single-occasion to serial testing protocols. In the absence of an established protocol for confirmation testing after a multi-cancer test, we also only quantify exposure to unnecessary confirmation tests, and not the number or cost of these tests, which may vary in terms of their invasiveness and accuracy. These features, as well as their implications for patient quality of life should be considered in a full accounting of the burden of unnecessary confirmation testing.

Our results are subject to limitations that stem mostly from the inputs used. First, diagnostic performance estimates were sourced from a published multi-cancer testing study.^3^ These performance estimates were not prospective; they were derived from patients with and without a diagnosed cancer. In a prospective setting of a healthy population, we expect lower sensitivity, particularly for early-stage tumors. Second, we approximated disease prevalence at the time of the test by the incidence within 5-year age groups and mortality by the risk of cancer-specific death over the next 15 years among individuals diagnosed at those ages. We acknowledge that, particularly for some cancers known or suspected to have longer latencies, the underlying prevalence at the time of the test could be higher than that assumed. Conversely, for cancers with shorter latencies and poorer baseline survival, the 15-year interval for baseline mortality might be too long. Using a shorter interval did not produce different decisions about which cancers to include, although the associated harm-benefit ratio was less favorable. Ideally, we would want to project the prevalence of early-stage cancer at the time of the test, but this would require additional data.

We used published cancer screening trials to provide a benchmark for how disease-specific mortality might be reduced by multi-cancer early detection. However, the extent to which these tests might prevent disease deaths is still highly uncertain and depends on how early they can reliably identify potentially fatal tumors as well as the efficacy of early treatment, which may vary across cancers. The mortality benefit also will be affected by how the tests are used in practice, whether alongside or instead of existing tests, such as those for breast, colorectal, and lung cancer. By utilizing current estimates of prevalence and mortality for these cancers, we modeled multi-cancer tests used alongside existing tests. This motivated our sensitivity analysis, which assumes a less pronounced mortality benefit among these cancers than among non-screened-for cancers.

While our analysis is designed to address primary questions about the population impact of multi-cancer testing, it also raises many more. Beyond metrics for harm and benefit and how to reliably approximate them, there are important questions about which cancers to include and how best to prioritize confirmation testing. A consensus about these matters will be needed before we can compare the different multi-cancer testing products currently under development. There are also important questions about the place of multi-cancer tests alongside established early detection modalities and how frequently the tests should be offered.

In conclusion, while emerging technology may facilitate detection of many cancers, its population impact depends on characteristics of both the cancers and the test. A key lesson from previous population screening tests is that the consequences of any early detection approach go far beyond test performance. Much more work is needed to determine how to deploy multi-cancer tests in a manner that optimizes impact and reduces the population cancer burden.

## Data Availability

No primary data has been used in this manuscript only published figures and public data summaries. All references and links to the published material and the public data resource are in the citations

## Notes

## Author contributions

Conceptualization, B.J. and R.E.; Methodology, B.J., R.G., and R.E.; Formal Analysis, B.J. and R.G.; Writing – Original Draft, B.J. and R.E.; Writing – Review & Editing, all authors; Visualization, R.G.

## Acknowledgements

We acknowledge helpful comments on previous drafts from Drs. Noel S. Weiss and Scott D. Ramsey.

## Data Availability

*The code for the model used in this article is available at* https://github.com/FredHutch/pancancer-testing-benefits-and-harms

