## Supplement for "A Quantitative Framework to Study Potential Benefits and Harms of Multi-cancer Early Detection Testing"

**Supplemental Appendix**

General expressions for the expected number of individual potentially exposed to unnecessary confirmation tests ($UCT$), expected number of cancers detected ($CD$), and expected number of lives saved ($LS$) for a $k$-cancer test are as follows:

$$UCT = N \cdot \left\{ \sum_{i=1}^{k} \rho_i \cdot P_i(T^+) \cdot [1 - L_i(T^+)] + \left(1 - \sum_{j=1}^{k} \rho_j \right) \cdot (1 - Sp) \right\} \quad (1)$$

$$CD = N \cdot \sum_{i=1}^{k} \rho_i \cdot P_i(T^+) \cdot L_i(T^+) \quad (2)$$

$$LS = N \cdot \sum_{i=1}^{k} r_i m_i \quad (3)$$

where $N$ is the number of individuals tested, $\rho_i$ is prevalence of cancer $i$, $P_i(T^+)$ is the overall sensitivity of the test for cancer $i$, $L_i(T^+)$ is the probability of correctly identifying cancer $i$, $Sp$ is the specificity of the test, $r_i$ is the mortality reduction associated with early detection of cancer $i$, and $m_i$ is the probability of death due to cancer $i$.

**Supplemental Table 1.** Hypothetical outcomes of a test for cancers A and B per 1,000 individuals: Expected number of individuals potentially exposed to unnecessary confirmation tests given sensitivity and specificity for both cancers and expected cancers detected. Results are shown for selected levels of prevalence of cancer B, overall sensitivity and localization probability and specificity for both cancers, and assuming the prevalence of cancer A is 0.1%.

| Sensitivity, % | Localization, % | Specificity, % | Prevalence, % | Unnecessary confirmation tests, n | Cancers detected, n |
|---|---|---|---|---|---|
| 50 | 50 | 95 | 0.05 | 50.3 | 0.4 |
| | | | 0.10 | 50.4 | 0.5 |
| | | | 0.50 | 51.2 | 1.5 |
| | | | 1.00 | 52.2 | 2.8 |
| 80 | 50 | 95 | 0.05 | 50.5 | 0.6 |
| | | | 0.10 | 50.7 | 0.8 |
| | | | 0.50 | 52.1 | 2.4 |
| | | | 1.00 | 53.9 | 4.4 |
| 50 | 80 | 95 | 0.05 | 50.1 | 0.6 |
| | | | 0.10 | 50.1 | 0.8 |
| | | | 0.50 | 50.3 | 2.4 |
| | | | 1.00 | 50.6 | 4.4 |
| 80 | 80 | 95 | 0.05 | 50.2 | 1.0 |
| | | | 0.10 | 50.2 | 1.3 |
| | | | 0.50 | 50.7 | 3.8 |
| | | | 1.00 | 51.2 | 7.0 |
| 50 | 50 | 99 | 0.05 | 10.4 | 0.4 |
| | | | 0.10 | 10.5 | 0.5 |
| | | | 0.50 | 11.4 | 1.5 |
| | | | 1.00 | 12.6 | 2.8 |
| 80 | 50 | 99 | 0.05 | 10.6 | 0.6 |
| | | | 0.10 | 10.8 | 0.8 |
| | | | 0.50 | 12.3 | 2.4 |
| | | | 1.00 | 14.3 | 4.4 |
| 50 | 80 | 99 | 0.05 | 10.1 | 0.6 |
| | | | 0.10 | 10.2 | 0.8 |
| | | | 0.50 | 10.5 | 2.4 |
| | | | 1.00 | 11.0 | 4.4 |
| 80 | 80 | 99 | 0.05 | 10.2 | 1.0 |
| | | | 0.10 | 10.3 | 1.3 |
| | | | 0.50 | 10.9 | 3.8 |
| | | | 1.00 | 11.7 | 7.0 |

**Supplemental Table 2.** Unnecessary confirmation tests, cancers detected, and lives saved associated with adding specific cancers to an existing test for breast cancer at specific screening ages assuming specificity of the two-cancer test is 99% and the mortality reduction for each cancer is 5% or 20%.

| Screening age, y | Breast + | Unnecessary confirmation tests, n | Cancers detected, n | Lives saved, n | |
|---|---|---|---|---|---|
| | | | | 5% reduction | 20% reduction |
| 50 | Lung | 10.1 | 1.8 | 0.1 | 0.2 |
| | Colorectal | 10.0 | 1.9 | 0.0 | 0.2 |
| | Ovary | 10.0 | 1.7 | 0.0 | 0.2 |
| | Pancreas | 10.1 | 1.6 | 0.0 | 0.1 |
| | Liver | 10.0 | 1.5 | 0.0 | 0.1 |
| 60 | Lung | 10.1 | 3.2 | 0.1 | 0.5 |
| | Colorectal | 10.1 | 3.1 | 0.1 | 0.4 |
| | Ovary | 10.1 | 2.6 | 0.1 | 0.3 |
| | Pancreas | 10.1 | 2.5 | 0.1 | 0.3 |
| | Liver | 10.1 | 2.4 | 0.1 | 0.3 |
| 70 | Lung | 10.2 | 4.4 | 0.3 | 1.1 |
| | Colorectal | 10.1 | 4.2 | 0.2 | 0.8 |
| | Ovary | 10.1 | 3.1 | 0.2 | 0.6 |
| | Pancreas | 10.1 | 3.1 | 0.2 | 0.6 |
| | Liver | 10.1 | 2.8 | 0.1 | 0.5 |

**Supplemental Table 3.** Unnecessary confirmation tests, cancers detected, and lives saved associated with adding specific cancers to an existing test for breast and lung cancers at specific screening ages assuming specificity of the three-cancer test is 99% and the mortality reduction for each cancer is 5% or 20%.

| Screening age, y | Breast+Lung+ | Unnecessary confirmation tests, n | Cancers detected, n | Lives saved, n | |
|---|---|---|---|---|---|
| | | | | 5% reduction | 20% reduction |
| 50 | Colorectal | 10.1 | 2.1 | 0.1 | 0.2 |
| | Ovary | 10.1 | 1.9 | 0.1 | 0.2 |
| | Pancreas | 10.1 | 1.8 | 0.1 | 0.2 |
| | Liver | 10.1 | 1.8 | 0.1 | 0.2 |
| 60 | Colorectal | 10.1 | 4.0 | 0.2 | 0.7 |
| | Ovary | 10.1 | 3.5 | 0.2 | 0.6 |
| | Pancreas | 10.2 | 3.4 | 0.1 | 0.6 |
| | Liver | 10.1 | 3.3 | 0.1 | 0.6 |
| 70 | Colorectal | 10.2 | 5.9 | 0.3 | 1.3 |
| | Ovary | 10.2 | 4.7 | 0.3 | 1.1 |
| | Pancreas | 10.3 | 4.7 | 0.3 | 1.2 |
| | Liver | 10.2 | 4.5 | 0.3 | 1.1 |

**Supplemental Figure 1.** Sensitivity analysis 1: Unnecessary confirmation tests per life saved associated with adding specific cancers to an existing test for breast cancer at specific screening ages (columns) assuming specificity of the two-cancer test is 99%, 15-year incidence-based mortality from the Surveillance, Epidemiology, and End Results program, and the mortality reduction for high-prevalence cancers is 10% while the mortality reduction for low-prevalence cancers is 50%.

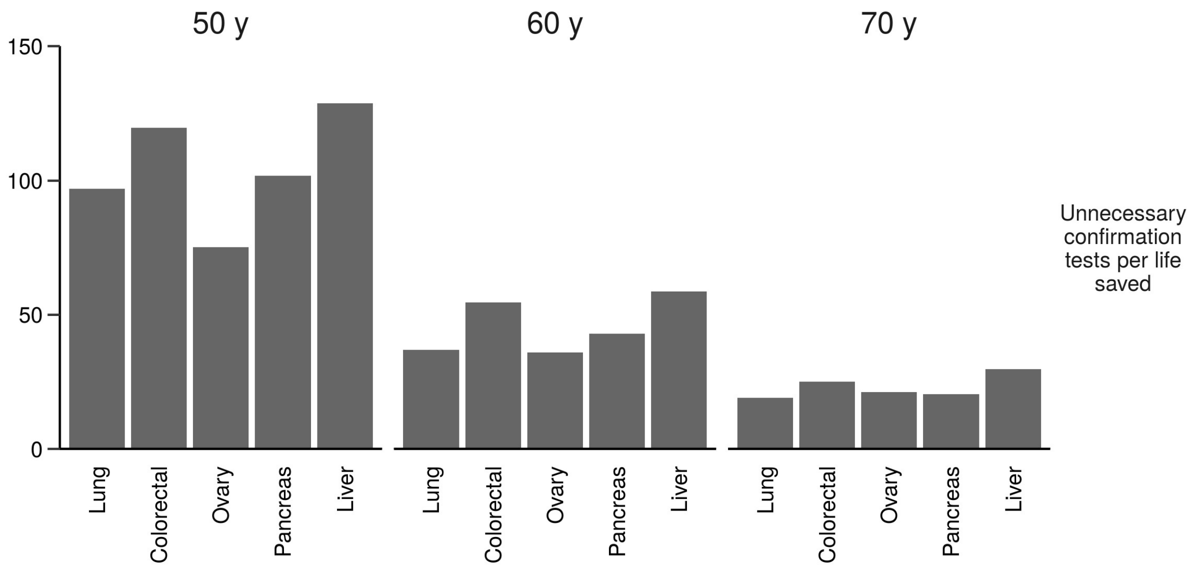

**Supplemental Figure 2.** Sensitivity analysis 2: Unnecessary confirmation tests per life saved associated with adding specific cancers to an existing test for breast cancer at specific screening ages (columns) assuming specificity of the two-cancer test is 99%, 10-year incidence-based mortality from the Surveillance, Epidemiology, and End Results program, and the mortality reduction for each cancer is 20%.

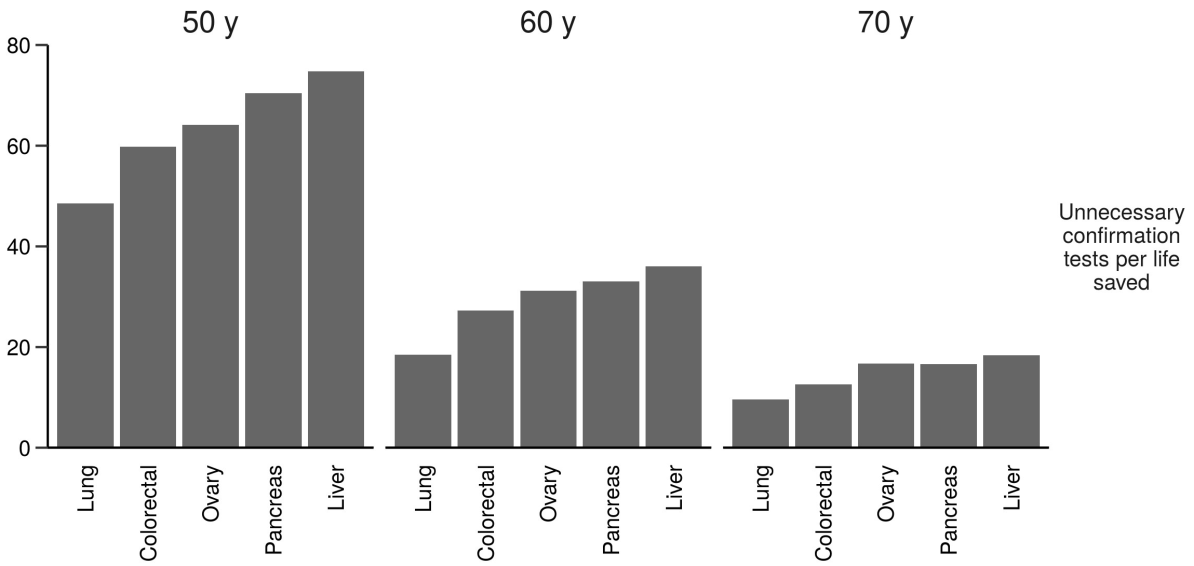
